# Social inequalities in the misbelief of chloroquine’s protective effect against COVID-19: results from the EPICOVID-19 study in Brazil

**DOI:** 10.1101/2023.05.29.23290677

**Authors:** Bruno P Nunes, Inácio Crochemore-Silva, Grégore I Mielke, Luis Paulo Vidaletti, Mariangela Freitas da Silveira, Pedro C Hallal

**Affiliations:** Postgraduate Programme in Nursing, Universidade Federal de Pelotas, Pelotas, Brazil; Postgraduate Programme in Epidemiology, Universidade Federal de Pelotas, Pelotas, Brazil; Postgraduate Programme in Physical Activity, Universidade Federal de Pelotas, Pelotas, Brazil; School of Public Health, The University of Queensland, Brisbane, Australia; College of Applied Health Sciences, Department of Kinesiology and Community Health, University of Illinois

**Keywords:** COVID-19. Chloroquine. Socioeconomic Factors. Epidemiology.

## Abstract

**Objectives:** The aim of this study was to assess the spread of denialist messages regarding COVID-19 in Brazil, specifically examining how social inequalities contributed to the misconception of chloroquine having a protective effect against the virus.

**Study design:** Three countrywide population-based studies were conducted in 2020 (May 14-21, June 4-7, and June 21-24), including 133 Brazilian cities (n=88,772).

**Methods:** Participants were asked whether they believed in chloroquine’s protective effect against infection with the SARS-CoV-2 virus (no/yes/don’t know). A jeopardy index score to assess cumulative social deprivation was calculated based on gender, racial and socioeconomic variables. Descriptive analysis and inequality measures (Slope Index of Inequality – SII; and Concentration Index – CIX) were used to evaluate the main association under investigation. Multinomial logistic regression was used to evaluate 3-category outcome according to independent variables.

**Results:** Overall, 47.9% of participants either believed that chloroquine prevented against COVID-19 or said, “I don’t know”. Misbelief and lack of knowledge about chloroquine were greater among the most vulnerable (lowest levels of education and socioeconomic status). Absolute and relative inequalities were observed according to jeopardy index. Lack of knowledge was 2.49 greater among women than among men. Race/ethnicity minorities, those with low education and low socioeconomic status were more likely to erroneously believe that chloroquine prevented against COVID-19. The highest absolute inequality was observed for the category “I don’t know” (SII = -14.3).

**Conclusions:** Misbelief of chloroquine’s protective effect against the SARS-CoV-2 virus was high in Brazil. People with greater social vulnerability were more likely to wrongly believe chloroquine prevented against COVID-19

## Introduction

The COVID-19 pandemic has caused several negative consequences for the world population^1, 2^. The most striking impact was the high avoidable mortality caused from inadequate management of the SARS-CoV-2 virus spread. Brazil holds the second-highest number of confirmed COVID-19 deaths (702,421; May 2023), with a mortality rate of over 330 deaths per 100,000 inhabitants. Despite representing 2.7% of the world population, the country accounts for 10.5% of the COVID-19 mortality as of September 2022^3^. In addition, mortality was unequally distributed in the population^4–6^. The higher mortality among most vulnerable individuals also worsened the ability of families to maintain and guarantee a family budget sufficient for dignified survival, increasing, for example, food insecurity^7^. In addition to population health issues, there was a deepening of social inequalities^8, 9^.

Inadequate pandemic management is related to scientific denialism^10^, as observed in Brazil^11, 12^. Throughout the pandemic, the former Brazilian president Jair Bolsonaro (mandate 2019-2022) carried out actions against science, such as political-party disputes with state governors, relativization of the magnitude and severity of the SARS-CoV-2 virus and creating a dichotomy between public health and economics. Even more severe was the strategy of denying scientific consensus, the delay in purchasing vaccines and the defense of medicines known to be ineffective for preventing and treating infection, such as chloroquine^13^.

As a result, there was a strong emphasis on chloroquine as an effective treatment of SARS-CoV-2 infection, leading to an indiscriminate use prescription by doctors. Unfortunately, the Federal Council of Medicine failed to fight chloroquine off-label use for COVID-19. Such strategies and actions influenced the population’s perception of the protective effect of chloroquine in preventing and treating SARS-CoV-2 infection^11^.

There is an assumption that scientific denialism related to chloroquine reached more the population that supports or is ideologically and electorally aligned with the President elected in 2018, which is characterized by men, older people, and individuals with greater social privileges (higher income and education, for example)^14^. Nevertheless, the complexity of information dissemination in society suggests that the fake news’ effect may not be restricted to electorally aligned people^4^. Unequal access to high-quality information, lower access to health services^15^, and greater difficulties for daily life can make the effect of the spread of fake news about chloroquine more frequent in the most vulnerable people.

In this context, intersectionality is a relevant way of evaluating health indicators to measure social inequities^16–19^. Previous evidence indicates more significant SARS-CoV-2 virus infection and mortality in contexts characterized by inequalities ^4, 5, 20^, especially among black, brown, or indigenous individuals and those presenting lower education and lower household assets levels^5, 6, 21, 22^. The systemic effect of scientific denialism in Brazil seems to widespread impact the society and not only on supporters of the current President, affecting the most vulnerable population deeply as found for mortality^4, 23^. Also, opinion surveys indicate that the positive perception of chloroquine is more concentrated among people with lower education level^24, 25^.

The aim of this study was to assess the spread of denialist messages regarding COVID-19 in Brazil, specifically examining how social inequalities contributed to the misconception of chloroquine having a protective effect against the virus. We hypothesized that scientific denialism is more concentrated in people with greater social vulnerability.

## Methods

Data are from the EPICOVID-19 Brazil study. These include three repeated seroprevalence studies conducted in 2020: 1) May 14 to 21 (n = 25,025); 2) June 4th to 7th (n = 31,165); and 3) June 21 to 24 (n = 33,207). For each face-to-face survey, a three-stage probabilistic sample was selected (cities, urban census tracts and households). Participants tested for antibodies against SARS-CoV-2 using the WONDFO SARS-CoV-2 Antibody Test (Wondfo Biotech Co., Guangzhou, China), which detects the presence of antibodies against SARS-CoV-2 (IgG and IgM). Further methodological details are available elsewhere^21, 26, 27^. For the present study, 88,772 individuals with valid information for beliefs regarding COVID-19 were included in the analysis.

The primary outcome assessed in this study was the percentage of the population holding the misconception that chloroquine provides protection against the SARS-CoV-2 virus. This was measured using the following question: “What do you believe offers protection against the coronavirus? [take chloroquine – no/yes/do not know”]. In our study, responses “yes” and “I do not know” were considered as an indicator of denialism in the pandemic due to the wide dissemination led by the actions and speeches of the President, using fake news and misinformation about the “efficacy” of chloroquine to prevent and treat COVID-19.

A jeopardy index score was calculated based on the aggregation of four sociodemographic variables that express dimensions of social privileges: gender (male; female), self-reported race/skin color (“white”; “brown”/mixed race; “black”; “yellow”; and “indigenous”), education level (incomplete elementary school; complete elementary school; complete high school; complete university degree); and wealth score (divided in quartiles), which was based on characteristics and assets of the household^28^. A composite jeopardy index score was created by assigning the most privileged group of each variable a score of zero (men, white, highest education, and highest socioeconomic position) and the least privileged group a score of one (women and non-white) or three (none or incomplete primary level of education and the lowest quartile of socioeconomic position). Therefore, for each variable the following scores were assigned: gender (men = 0; women = 1); racial identity (white = 0; non-white = 1); education (university graduate = 0; complete secondary or incomplete university = 1; complete primary or incomplete secondary = 2; none or incomplete primary = 3); socioeconomic position (top quartile = 0; 3rd quartile = 1; 2nd quartile = 2, bottom quartile = 3). Scores for each indicator were summed, resulting a ‘Jeopardy Index’ ranging from 0 to 8. The index is lower for individuals with greater social privilege (or greater guarantee of human rights), which can also be interpreted as lower social vulnerability.

We performed descriptive analysis using prevalence and respective confidence intervals (95%CI). The main outcome (“believe in the protective effect of chloroquine against the SARS-CoV-2 virus”) was analyzed according to the main exposure (jeopardy index) and by all variables that composed the index. We used multinomial logistic regression models to evaluate the crude and adjusted odds ratios of belief (yes) and lack of knowledge (don’t know) in the chloroquine protective effect against COVID-19 according to each independent variable (sex, skin color, education level and wealth) where all variables were mutually included in adjusted model. According to jeopardy index, we evaluate the crude effect of the belief (yes) and lack of knowledge (don’t know) in the chloroquine protective effect against COVID-19.

Complex measures of inequality were performed by calculating the Slope Index of Inequality (SII) and Concentration Index (CIX) using the jeopardy index as exposure. The SII represents the outcome’s absolute difference (in predicted values through logistic regression) according to the jeopardy index. Thus, the SII represents the absolute difference, in percentage points, between the values estimated for the extreme groups of the stratification variable. The linearity deviation was evaluated through visual assessment. The CIX evaluates the relative inequality similarly to the GINI index. Both indicators considered the entire distribution of the stratifier (jeopardy index) and were presented at values between -100 to +100. Negative and positive values express more pronounced inequalities among most and lowest vulnerable people, respectively^29, 30^.

## Results

Sociodemographic characteristics of the 88,772 eligible participants are described in Table 1. Most participants were women (58.3%), self-reported their skin color as brown, and completed high school. Not knowing or believing that chloroquine has a protective effect against COVID-19 was observed in almost half of the sample (47.9%). Overall, 20.0% (95%CI: 19.7; 20.4) of participants reported they believed that chloroquine was effective, while 27.9% (95%CI: 27.4; 28.3) reported they did not know.

**Table 1.** Sociodemographic characteristics and description and belief in the chloroquine protective effect against COVID-19 according to sociodemographic variables. Brazil, EPICOVID (rounds 1 to 3), 2020. N=88,772*

| Variables | % | Belief in chloroquine protective effect |  |  |
| --- | --- | --- | --- | --- |
|  |  | No<br>% (95%CI) | Yes<br>% (95%CI) | Don't know<br>% (95%CI) |
| Sex |  |  |  |  |
| Male | 41.7 | 51.4 (50.7; 52.0) | 22.0 (21.5; 22.5) | 26.6 (26.0; 27.2) |
| Female | 58.3 | 52.6 (52.1; 53.2) | 18.6 (18.2; 19.0) | 28.8 (28.2; 29.3) |
| Race/skin color |  |  |  |  |
| White | 37.0 | 53.7 (53.0; 54.4) | 18.5 (18.0; 19.0) | 27.8 (27.1; 28.5) |
| Brown (mixed race) | 45.9 | 51.5 (50.9; 52.2) | 21.4 (20.9; 21.9) | 27.1 (26.5; 27.7) |
| Black | 12.9 | 53.5 (52.4; 54.6) | 18.9 (18.1; 19.7) | 27.6 (26.7; 28.5) |
| Yellow | 2.8 | 50.2 (48.3; 52.0) | 21.3 (19.7; 23.0) | 28.5; 26.7; 30.4) |
| Indigenous | 1.4 | 46.5 (43.5; 49.5) | 24.5 (21.9; 27.3) | 29.0 (26.3; 31.8) |
| Education level |  |  |  |  |
| University degree | 20.1 | 63.9 (63.1; 64.7) | 16.5 (15.9; 17.2) | 19.5 (18.9; 20.2) |
| High school | 36.6 | 53.3 (52.7; 54.0) | 20.5 (20.0; 21.1) | 26.1 (25.5; 26.8) |
| Elementary school | 19.6 | 46.3 (45.4; 47.2) | 20.9 (20.2; 21.6) | 32.8 (32.0; 33.6) |
| Incomplete elementary school | 23.7 | 45.3 (44.4; 46.2) | 21.4 (20.7; 22.0) | 33.3 (32.4; 34.2) |
| Wealth quartiles |  |  |  |  |
| 1 (Richest) | 22.6 | 58.0 (57.3; 58.7) | 19.1 (18.5; 19.6) | 22.9 (22.3; 23.6) |
| 2<sup>nd</sup> | 23.8 | 54.0 (53.2; 54.9) | 19.0 (18.4; 19.6) | 27.0 (26.3; 27.7) |
| 3<sup>rd</sup> | 24.6 | 51.2 (50.5; 51.9) | 19.5 (19.0; 20.1) | 29.3 (28.5; 30.0) |
| 4 (Poorest) | 29.0 | 46.8 (46.0; 47.6) | 22.0 (21.4; 22.6) | 31.2 (30.5; 32.0) |
| **Total** | **100.0** | **52.1 (51.6; 52.6)** | **20.0 (19.7; 20.4)** | **27.9 (27.3; 28.3)** |

\*Analytical sample: valid information to variable "chloroquine protective effect against COVID-19".

The highest prevalence of lack of knowledge and belief in the protective effect of chloroquine was observed in participants who declared themselves to be indigenous. Participants with university degree had a lower percentage of belief in the effect of chloroquine (16.5%) than those with lower levels of education. A gradient in education levels and wealth quartiles among those who are unaware of the actual effect of chloroquine was observed. The prevalence of lack of knowledge regarding chloroquine’s protective effect was higher among individuals with lower education levels and those in the lowest wealth quartile. Conversely, among people who believe in the protective effect of chloroquine, the highest prevalence was observed among the lowest socioeconomic position group.

The associations of sociodemographic variables with lack of knowledge and the belief in the protective effect of chloroquine against COVID-19 are presented in Table 2. Women had lower odds of believing in the protective effects of chloroquine than men. Race/ethnicity minorities, as well as those with lower education levels and lower socioeconomic positions, had higher odds likely to erroneously believe that chloroquine prevented against COVID-19. Overall, women also and those with lower education and socioeconomic position were more likely than their counterpart to report lack of knowledge about the protective effects of chloroquine.

**Table 2.** Association between sociodemographic characteristics and misbelief in the protective effect of chloroquine against COVID-19. Brazil, EPICOVID (rounds 1 to 3), 2020. (n=88,772)

| Variables | Chloroquine protective effect<br>(Reference: No) <sup>a</sup> |  |  |  |
| --- | --- | --- | --- | --- |
|  | Crude |  | Adjusted <sup>b</sup> |  |
|  | Yes<br>OR (95%CI) | Don't know<br>OR (95%CI) | Yes<br>OR (95%CI) | Don't know<br>OR (95%CI) |
| **Sex** |  |  |  |  |
| Male | 1.00 | 1.00 | 1.00 | 1.00 |
| Female | 0.82<br>(0.79; 0.86) | 1.06<br>(1.02; 1.09) | 0.83<br>(0.80; 0.86) | 1.06<br>(1.03; 1.10) |
| **Race/skin color** |  |  |  |  |
| White | 1.00 | 1.00 | 1.00 | 1.00 |
| Non-white <sup>c</sup> | 1.17<br>(1.13; 1.22) | 1.02<br>(0.98; 1.06) | 1.09<br>(1.05; 1.14) | 0.92<br>(0.88; 0.95) |
| **Education level** |  |  |  |  |
| University degree |  |  |  |  |
| High school | 1.49<br>(1.41; 1.57) | 1.61<br>(1.53; 1.69) | 1.42<br>(1.34; 1.50) | 1.54<br>(1.46; 1.62) |
| Elementary school | 1.75<br>(1.65; 1.85) | 2.32<br>(2.20; 2.44) | 1.62<br>(1.52; 1.73) | 2.17<br>(2.06; 2.30) |
| Incomplete elementary school | 1.82<br>(1.71; 1.94) | 2.41<br>(2.27; 2.54) | 1.65<br>(1.54; 1.77) | 2.18<br>(2.04; 2.32) |
| **Wealth quartiles** |  |  |  |  |
| 1<sup>st</sup> (wealthiest) | 1.00 | 1.00 | 1.00 | 1.00 |
| 2<sup>nd</sup> | 1.07<br>(1.02; 1.13) | 1.26<br>(1.20; 1.33) | 0.97<br>(0.92; 1.03) | 1.12<br>(1.06; 1.18) |
| 3<sup>rd</sup> | 1.16<br>(1.11; 1.22) | 1.45<br>(1.38; 1.52) | 0.99<br>(0.94; 1.04) | 1.16<br>(1.10; 1.22) |
| 4<sup>th</sup> (Poorest) | 1.43<br>(1.36; 1.51) | 1.69<br>(1.61; 1.77) | 1.15<br>(1.09; 1.22) | 1.24<br>(1.17; 1.30) |

<sup>a</sup> Multinomial logistic regression model. Odds ratios indicate the odds of reporting “yes” or “I don’t know” instead of “No”. OR <1 indicates that the participants were less likely, and OR >1 suggests that they were more likely to respond “Yes” or “I don’t know”, instead of “No”, to the question “Do you believe chloroquine offers protection against the coronavirus?”
<sup>b</sup> Mutually adjusted for other variables presented in the table.
<sup>c</sup> Black-Brown-Yellow-Indigenous

The prevalence of belief and lack of knowledge in the protective effect of chloroquine against COVID-19 according to the Jeopardy index is presented in Figure 1. Across all groups, the majority of participants did not believe in the protective effect of chloroquine. However, as the Jeopardy index scores increased, the proportion of participants who did not believe in the protective effect of chloroquine decreased. Conversely, the percentage of people who were unaware of the effect of chloroquine increased as the Jeopardy index scores rose. In general, around 20% of people believed in the protective effect of chloroquine, and this percentage did not vary based on the Jeopardy index score. Compared with those in the lowest jeopardy index score [male, white, highest education, and highest wealth quartile]), those with a score of four onward had higher odds of reporting they believed in the protective effects of chloroquine (Table 3), while participants with a score of two or higher had higher odds of being unaware of the effect of chloroquine. Differences between scores were greater among those who are unaware of the effect of chloroquine compared to believers, summarized in the indicators of absolute and relative inequalities (SII and CIX, respectively).

**Figure 1.**
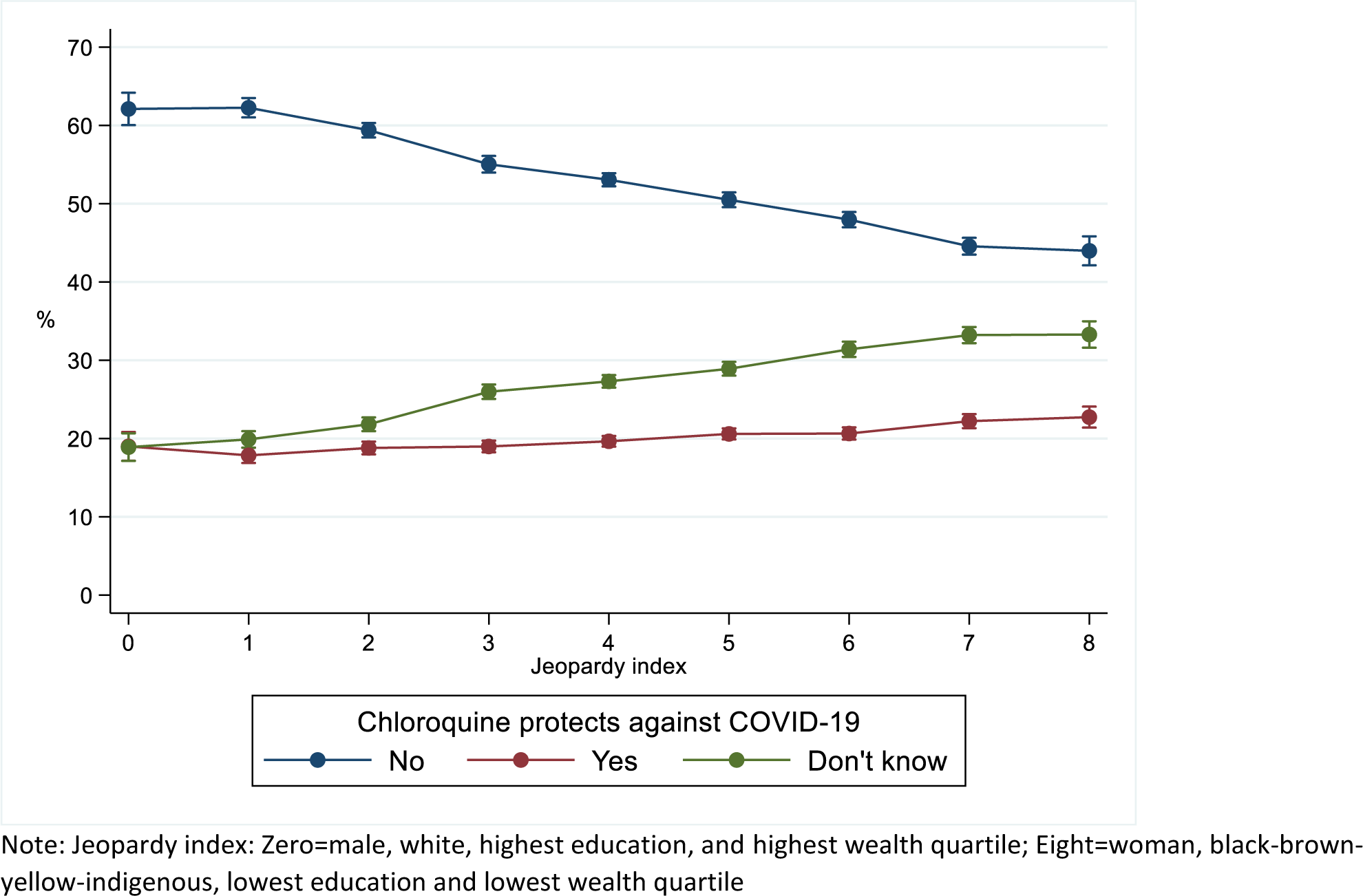
Predicted prevalence of the belief in the chloroquine protective effect against COVID-19 according to jeopardy index. Brazil, EPICOVID (rounds 1 to 3), 2020.

**Table 3.** Belief in the chloroquine protective effect against COVID-19 according to jeopardy index. Brazil, EPICOVID (rounds 1 to 3), 2020.

| Variables | Chloroquine protective effect<br>(Reference: No) <sup>a</sup> |  |
| --- | --- | --- |
|  | Yes<br>OR (CI95%) | Don't know<br>OR (CI95%) |
| Jeopardy index <sup>b</sup> |  |  |
| 0 | Ref | Ref |
| 1 | 0.94 (0.81; 1.08) | 1.05 (0.91; 1.21) |
| 2 | 1.03 (0.91; 1.18) | 1.21 (1.06; 1.38) |
| 3 | 1.13 (0.99; 1.28) | 1.55 (1.37; 1.76) |
| 4 | 1.21 (1.07; 1.37) | 1.69 (1.50; 1.91) |
| 5 | 1.33 (1.17; 1.52) | 1.88 (1.67; 2.12) |
| 6 | 1.41 (1.24; 1.60) | 2.15 (1.90; 2.43) |
| 7 | 1.63 (1.43; 1.86) | 2.45 (2.16; 2.78) |
| 8 | 1.69 (1.47; 1.94) | 2.49 (2.15; 2.87) |
| SII (95%CI) | -4.3 (-5.3; -3.3) | -14.3 (-15.5; -13.2) |
| CIX (95%CI) | -3.0 (-3.8; -2.3) | -6.6 (-7.2; -5.9) |

<sup>a</sup> Multinomial logistic regression model. Odds ratios indicate the odds of reporting “yes” or “I don’t know” instead of “No”. OR <1 indicates that the participants were less likely, and OR >1 suggests that they were more likely to respond “Yes” or “I don’t know”, instead of “No”, to the question “Do you believe chloroquine offers protection against the coronavirus?”
<sup>b</sup> Jeopardy index: Zero=male, white, highest education, and highest wealth quartile; Eight=woman, black-brown-yellow-indigenous, lowest education and lowest wealth quartile
<sup>c</sup> Slope index of inequality and Concentration Index (values between -100 a 100) according to jeopardy index.

## Discussion

The findings indicate that during the first months of the pandemic there was a very high misbelief in chloroquine’s protective effect against the SARS-CoV-2 virus. One in five Brazilians believed in the use of chloroquine as a protection against the coronavirus, while nearly a quarter were uncertain about its effects. Furthermore, our study showed notable disparities, with a higher prevalence of misbelief, particularly among the most vulnerable population groups. On the other hand, the belief in the protective effect of mask usage and adherence to stay-at-home policies were nearly universal and did not exhibit inequalities based on the jeopardy index (data not shown). These findings demonstrate the specific sources of denialism in Brazil and their influence on public perceptions.

The high misbelief in the protective effect of chloroquine against the COVID-19 could be attributed to the data collection period, which was carried out between May and August 2020. However, it is important to note that there was a high assertive belief in the protective effect of chloroquine (20% answered “yes”) even with results and scientific consensus described in the first half of 2020 already pointing out the lack of robust evidence on the benefit of chloroquine and warned of its side effects. Since April 2020, the preliminary results of the CloroCovid-19 study pointed out the inefficacy and the risks of chloroquine in the disease treatment^33^.

In addition, the most current evidence indicates a similar percentage of the Brazilian population that continues to believe in the protective effect of chloroquine. The survey “The Face of Democracy”, for example, identified, in April 2021, that one in four Brazilians claims to have used drugs of the so-called “early treatment” against COVID-19 or to prevent infection by the SARS-CoV-2 virus.

The executive power’s milestone of the public defense of chloroquine/hydroxychloroquine happened on March 27, 2020, by a post of the President on a social network. The post cited “accurate information” about chloroquine having a “high success rate” using as a basis a pre-print paper (published only in July 2020) that evaluated the effect of hydroxychloroquine in a non-randomized study with 36 French patients. This post was in line with publications by the President of the United States at the time^34^. In February 2020, a parliamentary commission was created to deal with the COVID-19 crisis. In two sessions held in April and July 2020, chloroquine was the central meeting emphasis influenced by the speeches and posts of the head of the federal government. In these meetings, a parliamentarian aligned with the federal government informed the work of the Brazilian Army in manufacturing the drug to be sent to states and municipalities. Since then, technical documents and positions of the Federal Council of Medicine have been published supporting chloroquine mainly in the so-called “early treatment”, even for cases with mild symptoms, as observed in an Informative Note from May 2020. Since then, strategies such as the COVID Kit have been popularized, including using federal government digital platforms (*TrateCOV*)^34^. In this context, more robust scientific evidence has been signaling the null effect of chloroquine in the treatment and prevention of COVID-19, in addition to reports of the side effects which were widely recorded throughout the pandemic^35–37^.

The US Food and Drug Administration (FDA) stated, in June 2020, the ineffectiveness of chloroquine and hydroxychloroquine to treat SARS-CoV-2 infection. During the same period, the World Health Organization announced the interruption of clinical trials using chloroquine. In Brazil, except for the Federal Council of Medicine, scientific societies, scientists, and health councils warned about the risks of using chloroquine to face the pandemic. In May 2020, the National Health Council published a note indicating that scientific evidence pointed to side effects and no benefit for outcomes related to COVID-19^38^. Nevertheless, the defense of the drug by the president of the republic and his supporters continued throughout 2020 and 2021^13, 39^. Still, in 2022, statements about the benefit of chloroquine are found in several public interviews carried out by the head of the executive power^39^. It is also worth noting that in addition to the negationist discursive practice, there was a deliberate omission on the subject at the Ministry of Health level, executing imprecise positions or campaigns with adequate information to face the pandemic.

The ineffectiveness of chloroquine (or hydroxychloroquine) to combat the SARS-CoV-2 virus is more than consolidated^40–42^. Findings from the DETECTCOV-19 cohort in Manaus indicate a higher seroprevalence (immunoglobulin G positivity) among people who self-medicated as a prophylaxis strategy^43^. Similar results were observed in studies on the effect of hydroxychloroquine as a preventive or post-exposure therapy^44^. However, the population’s perception of chloroquine’s benefit (and others such as ivermectin) persists, according to results from 2021 and 2022^24, 45, 46^.

Scientific denialism as a government policy has caused a greater concentration of misperception about the role of chloroquine in facing the pandemic. Our findings identify inequalities, especially for those who did not know if the drug protected against the SARS-CoV-2 virus. Although the answer “I do not know” seems to be a lack of knowledge, it represents the population’s doubt influenced by the president’s speeches. It is noteworthy that for effective non-pharmacological measures to prevent infection (staying at home and wearing a mask), the percentage of respondents who did not know was <1%, respectively (data not shown). Thus, lack of knowledge seems to be more influenced by the government’s denialist strategy than by a scientific question about the drug’s effect. Thus, it is plausible to understand that the strategy of the Brazilian federal government was an initiative to discredit scientific consensus and create distrust in the population, making it challenging to adopt really effective strategies to face the pandemic^12, 47–50^. The perception of the available treatments tends to reduce the adoption of preventive measures. The governmental strategy aimed to herd immunity (i.e., “greater contamination possible”)^13, 51^ to achieve economic goals unrelated to basic human rights for life’s protection. The herd immunity proved impractical as was speculated by science since 2020, the chloroquine treatment was ineffective, and the result was high mortality attributable to the federal government’s denialist policy of the federal government^4, 52^.

In addition to study limitations related to the sampling and sample characteristics^21, 22^, a possible limitation of this analysis is related to the percentage of losses and refusals (46-47%), which can lead to a differential bias. The refusal might be higher among wealthier people who believe in the chloroquine effect^31, 32^. However, this possible bias may produce an underestimating misbelief in chloroquine’s protective effect against the SARS-CoV-2 virus. Thus, the possible selection bias may be overestimating the association, but simultaneously decreasing the magnitude of the misbelief in the Brazilian population’s chloroquine effect.

The misbelief in chloroquine’s protective effect was widespread in the Brazilian population, not just the profile of people ideologically and electorally aligned with the President of the Republic. People with greater social vulnerability were those most affected by a lack of knowledge about the effectiveness of chloroquine. The denialist federal management of the pandemic in Brazil was contaminant and contaminating, influencing the tragic mortality observed in the country.

### Statements of ethical approval

This study complied with all ethical precepts and legislation governing research with human beings and was approved by the National Research Ethics Commission (CAAE 30721520.7.1001.5313). All participants signed an informed consent form.

### Data sharing

The EPICOVID19 datasets are freely available online.

## Data Availability

The EPICOVID19 datasets are freely available online.

https://serrapilheira.org/pesquisadores/epicovid-19/

## Acknowledgments

The study was funded by the Brazilian Ministry of Health, Instituto Serrapilheira, Brazilian Collective Health Association and JBS Fazer o Bem Faz Bem. The funder has no role in study design, data collection/analysis, interpretation of findings, or manuscript writing.

